# Waning COVID-19 Vaccine Effectiveness for BNT162b2 and CoronaVac in Malaysia: An Observational Study

**DOI:** 10.1101/2022.01.15.22269326

**Authors:** Jing Lian Suah, Masliyana Husin, Peter Seah Keng Tok, Boon Hwa Tng, Thevesh Thevananthan, Ee Vien Low, Maheshwara Rao Appannan, Faizah Muhamad Zin, Shahanizan Mohd Zin, Hazlina Yahaya, Kalaiarasu M. Peariasamy, Sheamini Sivasampu

**Affiliations:** Institute for Clinical Research, National Institutes of Health, Ministry of Health Malaysia, Setia Alam 40170, Malaysia; Disease Control Division, Ministry of Health Malaysia, Putrajaya 62590, Malaysia; Medical Development Division, Ministry of Health Malaysia, Putrajaya 62590, Malaysia

**Author notes:** Corresponding Author(s): Jing Lian Suah, Institute for Clinical Research, National Institute of Health, Ministry of Health, Malaysia, No.1 Jalan Setia Murni U13/52 Seksyen U13, 40170 Shah Alam, Selangor. No. / ext: 8790, Masliyana Husin, Institute for Clinical Research, National Institute of Health, Ministry of Health, Malaysia, No.1 Jalan Setia Murni U13/52 Seksyen U13, 40170 Shah Alam, Selangor. No. / ext: 8816. Contributed equally to the manuscript (Jing Lian Suah, Masliyana Husin).

## Abstract

**Background:** Evaluation of vaccine effectiveness over time against severe acute respiratory syndrome coronavirus 2 (SARS-CoV-2) infection or coronavirus disease 2019 (COVID-19) is important. Evidence on effectiveness over time for the CoronaVac vaccine is lacking despite its widespread use globally. In Malaysia, a diverse set-up of COVID-19 vaccines was rolled out nationwide, and the waning of vaccine protection is a concern. We aimed to investigate and compare waning vaccine effectiveness against COVID-19 infections, COVID-19 related ICU admission and COVID-19 related deaths for BNT162b2 and CoronaVac vaccines.

**Methods:** In this observational study, we consolidated nationally representative data on COVID-19 vaccination and patients’ outcomes. Data on all confirmed COVID-19 cases from 1 to 30 September 2021 were used to compare vaccine effectiveness between the ‘early’ group (fully vaccinated in April to June 2021) and the ‘late’ group (fully vaccinated in Jul to Aug 2021). We used a negative binomial regression model to estimate vaccine effectiveness against COVID-19 infections for both ‘early’ and ‘late’ groups, by comparing the rates of infection for individuals vaccinated in the two different periods relative to the unvaccinated. Among confirmed COVID-19 cases, we used logistic regression to estimate and compare vaccine effectiveness against ICU admission and deaths between the two different periods.

**Findings:** For BNT162b2, vaccine effectiveness against COVID-19 infections declined from 90.8% (95% CI 89.4, 92.0) in the late group to 79.1% (95% CI 75.8, 81.9) in the late group. Vaccine effectiveness for BNT162b2 against ICU admission and deaths were comparable between the two different periods. For CoronaVac, vaccine effectiveness waned against COVID-19 infections from 74.4% in the late group (95% CI 209 70.4, 77.8) to 30.0% (95% CI 18.4, 39.9) in the early group. It also declined significantly against ICU admission, dropping from 56.1% (95% CI 51.4, 60.2) to 29.9% (95% CI 13.9, 43.0). For deaths, however, CoronaVac’s effectiveness did not wane after three to five months of full vaccination.

**Interpretation:** Vaccine effectiveness against COVID-19 infections waned after three to five months of full vaccination for both BNT162b2 and CoronaVac in Malaysia. Additionally, for CoronaVac, protection against ICU admission declined as well. Evidence on vaccine effectiveness over time informs evolving policy decisions on vaccination.

## INTRODUCTION

Since early 2021, clinical trials^1–3^ and real-world studies on vaccine effectiveness^4–6^ have demonstrated efficacy and effectiveness measures well above the World Health Organization (WHO)’s benchmark of 50%.^7^ This led to regulatory approval and widespread global use of an increasing number of vaccines. Vaccination also appeared to be substantially protective against severe disease from all main variants, including the Delta (B.1.617.2) variant.^5,8^ However, while booster dose(s) are being rolled out globally, uncertainty over the scale and pace in the waning of immunity underscores consideration over the timing for booster dose(s) in the future.

Concerns over possible waning of the protective immunity conferred by the vaccines emerged when breakthrough infections and disease were increasingly documented among fully vaccinated persons. In the USA, although still largely protective against hospital admissions, the effectiveness of the BNT162b2 vaccine against severe acute respiratory syndrome coronavirus 2 (SARS-CoV-2) infections declined from 88% during the first month after full vaccination to 47% after five months.^9^ Similar pattern of waning in the effectiveness of the BNT162b2 vaccine against SARS-CoV-2 infections was observed in Qatar^10^ and Israel.^11^ In the UK, vaccine effectiveness against symptomatic disease fell after 20 weeks against the Delta variant among both BNT162b2 and AZD1222 vaccine recipients.^12^ Evidence on the protection of COVID-19 vaccines against severe outcomes over time is mixed.^9–12^ Although immunogenicity studies showed that adaptive immune responses, such as memory B cells, remain in the circulation over time after full vaccination, findings were limited to six to eight weeks of follow-up.^13^ Moreover, evidence on the association between these adaptive responses and protection against severe disease and fatality in the real-world remains scarce, especially for the CoronaVac vaccine, despite its dominant use in many low to middle-income countries (LMICs).^14^

Malaysia’s nationwide mass COVID-19 immunisation rollout, the National COVID-19 Immunisation Programme (*Program Imunisasi COVID-19 Kebangsaan; PICK*), began on 24 February 2021. As of 31 August 2021, 65·4% of the adult population has been fully vaccinated.^15^ PICK utilised a diverse set-up of predominantly BNT162b2, CoronaVac, and AZD1222 vaccines, owing to global vaccine inequity and broadly lower supply to LMICs. This led to widespread use of CoronaVac, then approved for usage among individuals aged 18 years and older, including the elderly. CoronaVac accounted for 49·9% of all completed vaccinations as of 31 August 2021. The utilisation breakdowns of the three vaccines by phases of PICK are provided in Table S1 of the Supplementary Appendix. The chronology of PICK’s phases, and the effectiveness of the vaccines, were described previously.^6^ However, breakthrough COVID-19 cases started to increase since late July 2021, with a cumulative rate of 1·09% up to 31 August 2021. Additionally, breakthrough ICU admission and death rates were both close to 0·01% (Figures S1, S2 and S3 of the Supplementary Appendix). Hence, there is an imminent need to investigate whether the protective immunity wanes over time, which underpins policy decisions on the need, and timing, for booster dose(s) of COVID-19 vaccination. Given that Malaysia and other LMICs are faced with limited supply and potentially increased hesitancy, research on waning effectiveness will directly impact the dynamic planning and redistribution of boosters.

Given the lack of immunogenicity studies in Malaysia, our study is instead grounded on real-world data consolidated from nationally representative data on COVID-19 vaccination and patients’ outcomes. We investigated the presence and scale of waning vaccine effectiveness against COVID-19 infection, COVID-19-related ICU admission and COVID-19-related death for BNT162b2 and CoronaVac vaccines among adults in Malaysia.

## METHODS

### Data Environment

In Malaysia, the notification of COVID-19 cases and deaths are legally mandated under the provision of the Prevention and Control of Infectious Diseases Act 1988 (Act 342). At the time of writing, aggregated and granular data on COVID-19 cases, deaths, and vaccination are available and updated continuously on the Ministry of Health’s GitHub repository.^15,16^

Data of all confirmed COVID-19 cases were extracted from the Malaysia national electronic COVID-19 cases register, which is the national COVID-19 surveillance system. Using case and personal identification numbers, all confirmed COVID-19 cases were linked deterministically with their vaccination date and status (using the COVID-19 vaccine recipients line listing) and clinical outcomes of interest: admission into ICU (using the ICU admissions register) and deaths (using the COVID-19 deaths line listing). Details on the definition of outcomes and data sources have been described previously.^6^

### Study Design, and Comparator Groups

The study period spanned 1 September 2021 to 30 September 2021, during which the Delta variant was predominant in Malaysia.^17^ All individuals with confirmed SARS-CoV-2 infections occurring outside of this period or have received vaccines other than homologous CoronaVac and BNT162b2 were excluded. Although AZD1222 was the third most predominant vaccine in PICK, the first individual fully vaccinated with AZD1222 was on 21 July 2021, resulting in an insufficient follow-up period to evaluate the waning of vaccine effectiveness at the point of analysis. Full vaccination status is defined as ≥ 14 days after the receipt of the second dose of any of the two vaccines (BNT162b2 and CoronaVac) in this study.

Our study compared three groups – (i) those vaccinated in April to June 2021 (‘early’ group), corresponding to Phases one and two of PICK, (ii) those vaccinated in July to August 2021 (‘late’ group), corresponding to Phase three of PICK, and (iii) those unvaccinated in September 2021. This grouping enabled estimation of vaccine effectiveness at one to two months, and three to five months post-vaccination. Due to insufficient granularity on individual-level data for those who tested negative, we could not infer waning effectiveness as in a traditional cohort or case-control study, which includes uninfected individuals. Hence, we investigated whether the period of vaccination affected the rate of confirmed SARS-CoV-2 infection using a retrospective population cohort approach. We subsequently investigated the rates of COVID-19-related ICU admission and death using a retrospective cohort of confirmed COVID-19 cases, which exploited granular data to adjust for confounders.

### Retrospective Population Cohort using Census Data: Study Design, Methodology, and Statistical Analysis

We constructed a unified population cohort by merging data on vaccination, confirmed COVID-19 cases, and the Department of Statistics Malaysia’s Current Population Estimates^18^ for year 2021 to compare the rates of infection for individuals vaccinated in different periods relative to those unvaccinated. This analysis included individuals aged 15 and above, which reflects the age groups for which census-based population estimates were available, whereas estimates for the cut off of 18 years and above were unavailable.

We calculated the number of events for outcomes in the study period, and number of individuals vaccinated, indexed to the timing of vaccination (for those who were vaccinated), age groups, sex, and states of vaccination. We calculated the size of the unvaccinated population over time from the number of vaccinated individuals and the census data. Vaccine effectiveness by vaccine types for both ‘early’ and ‘late’ groups are estimated with a negative binomial regression, adjusting for (i) age groups, (ii) state or region, and (iii) sex, with the cumulative person-days as the offset, and the unvaccinated group as the baseline. Further details on the negative binomial regression model were described in the Supplementary Appendix.

### Retrospective Cohort of Confirmed COVID-19 Cases: Study Design, Methodology, and Statistical Analysis

Waning vaccine effectiveness against severe outcomes (ICU admission and death) over time was investigated by comparing the odds between individuals vaccinated in different periods relative to those unvaccinated, among confirmed COVID-19 cases.

The study population includes all confirmed COVID-19 cases aged 18 and above, with confirmatory test dates of between 1 September 2021 and 30 September 2021. To account for delays in the onset of severe outcomes post-confirmation of infection, the cohort records all deaths and ICU admissions occurring in September 2021 and up to November 2021. Vaccine effectiveness over time is estimated using a logistic regression, adjusting for (i) age as a continuous variable, (ii) presence of comorbidities, (iii) sex, (iv) nationality, and (v) state. Further details on the logistic regression model are in the Supplementary Appendix.

### Sensitivity Analysis

We conducted three sensitivity analyses. Firstly, we redefined ICU admissions and deaths as one outcome (‘severe’). Secondly, we stratified by age groups to evaluate age-specific waning of effectiveness. Thirdly, we inspected robustness to the inclusion and exclusion of all combinations of confounders in the logistic regression. All sensitivity analyses are reported in the Supplementary Appendix.

All analyses were conducted with Python, version 3.9, and R, version 4.1.2.

### RECoVaM study and Ethical Consideration

This study, commissioned by the Ministry of Health, Malaysia, is part of The Real-World Evaluation of COVID-19 Vaccines under the Malaysia National COVID-19 Immunisation Programme (RECoVaM) study registered in the National Medical Research Register (NMRR-21-1660-60697). This study was conducted according to guidelines in the Declaration of Helsinki and was granted ethnical approval by the Medical Research and Ethics Committee (MREC), Ministry of Health, Malaysia.

## RESULTS

### Retrospective Population Cohort using Census Data

Figure 1(a) depicts the eligible cohort of fully vaccinated individuals. After excluding individuals with infections outside of the study period and recipients of other vaccines, 9,927,350 individuals were fully vaccinated with homologous BNT162b2 or CoronaVac vaccines. There were 246,029 confirmed COVID-19 cases throughout the outcome observation period in September 2021, of which 163,902 were vaccinated, and 82,127 were unvaccinated.

**Figure 1:**
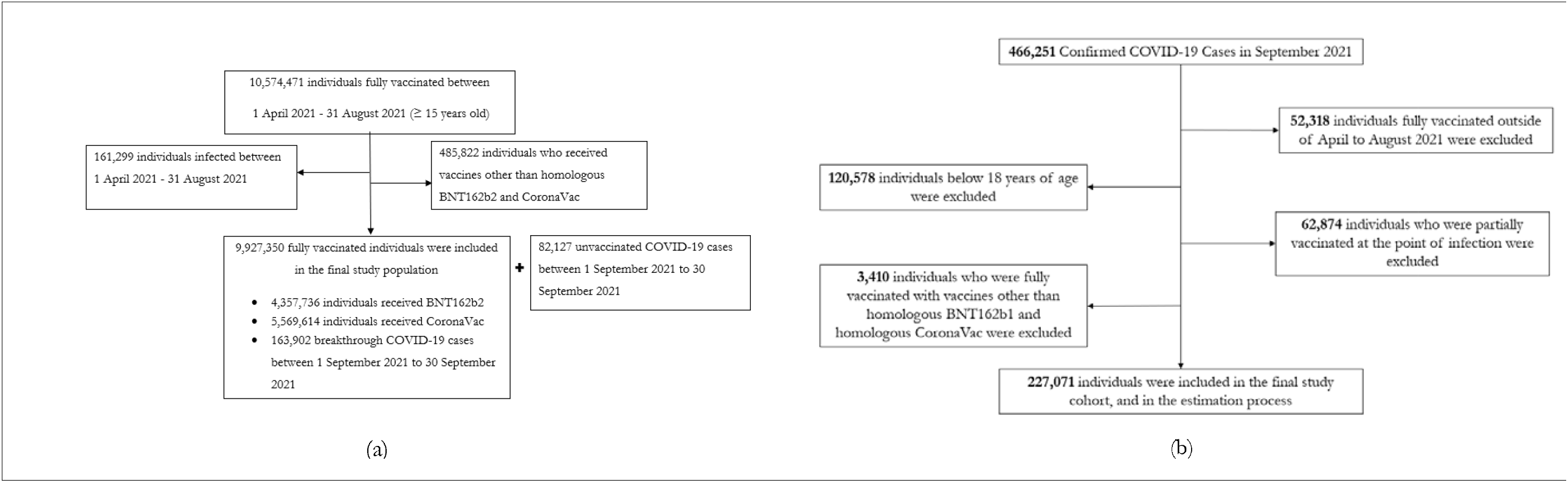
Study participants and cohort eligibility for the retrospective population cohort using census data, and the retrospective cohort of confirmed COVID-19 cases. (a) Individuals who were fully vaccinated with homologous CoronaVac or BNT162b2 between 1 April 2021 and 31 August 2021, aged 15 and above, had no documented COVID-19 infection prior to the outcomes observation period (1 September 2021 to 30 September 2021). (b) Confirmed COVID-19 cases between 1 September 2021 and 30 September 2021, aged 18 and above, with no prior documented COVID-19 infection, and fully vaccinated with homologous BNT162b2 and CoronaVac fully vaccinated (14 days post-dose two) between 1 April 2021 and 31 August 2021, or unvaccinated.

Table 1 presents the baseline characteristics by type of vaccine received and the timing of vaccination (‘early’ and ‘late’ groups) for the fully vaccinated individuals included in the cohort. Overall, there were more individuals fully vaccinated with BNT162b2 in the early group while the reverse is observed in the late group. This corresponds to the composition of vaccine platforms utilised across the various phases of PICK, as described previously.^6^

**Table 1.**
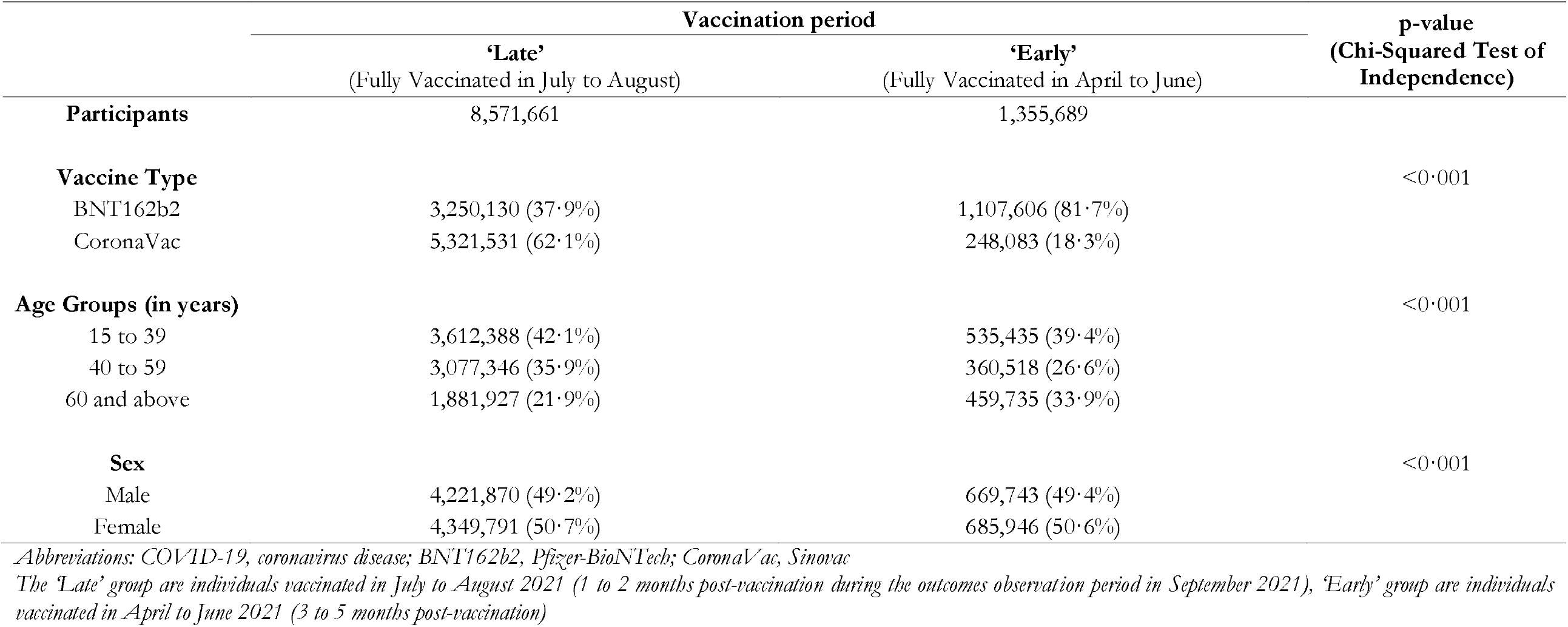
Baseline characteristics of the study population by duration since full vaccination.

|  | Vaccination period |  | p-value<br>(Chi-Squared Test of<br>Independence) |
| --- | --- | --- | --- |
|  | 'Late'<br>(Fully Vaccinated in July to August) | 'Early'<br>(Fully Vaccinated in April to June) |  |
| **Participants** | 8,571,661 | 1,355,689 |  |
| **Vaccine Type** |  |  | <0.001 |
| BNT162b2 | 3,250,130 (37.9%) | 1,107,606 (81.7%) |  |
| CoronaVac | 5,321,531 (62.1%) | 248,083 (18.3%) |  |
| **Age Groups (in years)** |  |  | <0.001 |
| 15 to 39 | 3,612,388 (42.1%) | 535,435 (39.4%) |  |
| 40 to 59 | 3,077,346 (35.9%) | 360,518 (26.6%) |  |
| 60 and above | 1,881,927 (21.9%) | 459,735 (33.9%) |  |
| **Sex** |  |  | <0.001 |
| Male | 4,221,870 (49.2%) | 669,743 (49.4%) |  |
| Female | 4,349,791 (50.7%) | 685,946 (50.6%) |  |

*Abbreviations: COVID-19, coronavirus disease; BNT162b2, Pfizer-BioNTech; CoronaVac, Sinovac*
*The 'Late' group are individuals vaccinated in July to August 2021 (1 to 2 months post-vaccination during the outcomes observation period in September 2021), 'Early' group are individuals vaccinated in April to June 2021 (3 to 5 months post-vaccination)*

Table 2 presents the unadjusted incidence rates for COVID-19 infection and vaccine effectiveness for both early and late groups among recipients of both vaccines (BNT162b2 and CoronaVac). Suppose there is no waning of effectiveness, one would expect to observe the similar incidence rates in both groups. However, for fully vaccinated individuals with either vaccine (BNT162b2 and CoronaVac), the incidence rates in the early group are higher than in the late group, indicating possible waning protection offered by the vaccines.

For BNT162b2, vaccine effectiveness against COVID-19 infection declined from 90·8% (95% CI 89·4, 92·0) among individuals vaccinated in the early group to 79·1% (95% CI 75·8, 81·9) in the late group. The degree of waning was observed to be greater among individuals in older age groups (aged 60 years and above, and 40 to 59 years old), when compared to their younger counterparts.

For CoronaVac, vaccine effectiveness against COVID-19 infection declined from 74·4% in the late group (95% CI 70·4, 77·8) to 30·0% (95% CI 18·4, 39·9) in the early group. Similar to the findings for BNT162b2, CoronaVac exhibits greater waning of effectiveness against COVID-19 infections among the older age groups.

**Table 2.** Vaccine effectiveness against COVID-19 infection by vaccine type and duration since full vaccination.

|  |  | 'Late'<br>(Fully Vaccinated in July to August) |  |  |  |  | 'Early'<br>(Fully Vaccinated in April to June) |  |  |  |  |
| --- | --- | --- | --- | --- | --- | --- | --- | --- | --- | --- | --- |
|  |  | Total | Person-days<br>at risk | Incidence<br>rates/1,000<br>persons | VE | CI (95%) | Total | Person-days<br>at risk | Incidence<br>rates/1,000<br>persons | VE | CI (95%) |
| **COVID-19 Infection** |  |  |  |  |  |  |  |  |  |  |  |
| **Overall** | Unvaccinated | 82,127 | 46,760,638 | 1.756 | ref | ref | 82,127 | 46,760,638 | 1.756 | ref | ref |
|  | BNT162b2 | 30,332 | 136,048,292 | 0.223 | 90.8 | 89.4, 92.0 | 22,105 | 41,577,287 | 0.532 | 79.1 | 75.8, 81.9 |
|  | CoronaVac | 105,397 | 141,117,636 | 0.747 | 74.4 | 70.4, 77.8 | 6,068 | 3,828,752 | 1.585 | 30.0 | 18.4, 39.9 |
| **Age Group**<br>**(in years)** |  |  |  |  |  |  |  |  |  |  |  |
| 15 to 39 | Unvaccinated | .. | .. | .. | ref | ref | .. | .. | .. | ref | ref |
|  | BNT162b2 | .. | .. | .. | 91.8 | 90.1, 93.3 | .. | .. | .. | 86.0 | 83.0, 88.4 |
|  | CoronaVac | .. | .. | .. | 73.6 | 68.0, 78.2 | .. | .. | .. | 66.9 | 59.9, 72.9 |
| 40 to 59 | Unvaccinated | .. | .. | .. | ref | ref | .. | .. | .. | ref | ref |
|  | BNT162b2 | .. | .. | .. | 88.8 | 86.4, 90.8 | .. | .. | .. | 76.9 | 71.9, 81.0 |
|  | CoronaVac | .. | .. | .. | 70.2 | 63.8, 75.4 | .. | .. | .. | 31.8 | 16.0, 44.6 |
| 60 and above | Unvaccinated | .. | .. | .. | ref | ref | .. | .. | .. | ref | ref |
|  | BNT162b2 | .. | .. | .. | 91 | 99.0, 99.4 | .. | .. | .. | 69.7 | 63.5, 74.8 |
|  | CoronaVac | .. | .. | .. | 78.6 | 74.4, 82.2 | .. | .. | .. | -38.2 | -67.9, -13.8 |

Abbreviations: CI, confidence intervals; COVID-19, coronavirus disease; VE, vaccine effectiveness; BNT162b2, Pfizer-BioNTech; CoronaVac, Sinovac.
The estimates were adjusted for age, gender and states of residence
The analysis compared between the 'Late' group, individuals vaccinated in July to August 2021 (1 to 2 months post-vaccination during the outcomes observation period in September 2021), and the 'Early' group, individuals vaccinated in April to June 2021 (3 to 5 months post-vaccination), and unvaccinated individuals by August 31, 2021, who act as the baseline

### Retrospective Cohort of Confirmed COVID-19 Cases

Vaccine effectiveness over time against ICU admission and death were measured using the adjusted odds ratio between different period of vaccination, relative to the unvaccinated, among confirmed COVID-19 cases in September 2021. Figure 1(b) shows the selection criteria for the study cohort, which includes 227,071 confirmed COVID-19 cases.

Table 3 presents the baseline characteristics for the 277,071 confirmed COVID-19 cases according to the status of vaccination, type of vaccination received (BNT162b2, CoronaVac) and the timing of vaccination. The crude numbers here, however, needs to be enumerated into incidence rates and adjusted for potential confounders, before vaccine effectiveness can be inferred.

**Table 3.** Baseline characteristics of the retrospective cohort of confirmed COVID-19 cases.

|  | Unvaccinated | BNT162b2 |  | CoronaVac |  | p-value<br>(Chi-Squared<br>Test of<br>Independence) |
| --- | --- | --- | --- | --- | --- | --- |
|  |  | ‘Late’<br>(Fully Vaccinated in<br>July to August) | ‘Early’<br>(Fully Vaccinated in<br>April to June) | ‘Late’<br>(Fully Vaccinated in<br>July to August) | ‘Early’<br>(Fully Vaccinated in<br>April to June) |  |
| **Participants** | 63,098 | 30,350 | 22,125 | 105,416 | 6,082 |  |
| **Admitted to ICU** | 1,242 (2.0%) | 102 (0.3%) | 91 (0.4%) | 834 (0.8%) | 110 (1.8%) | <0.001 |
| **Death** | 2,562 (4.1%) | 177 (0.6%) | 98 (0.4%) | 766 (0.7%) | 85 (1.4%) | <0.001 |
| **Age Groups (in years)** |  |  |  |  |  | <0.001 |
| 18 to 39 | 40,213 (63.7%) | 12,753 (42.0%) | 11,921 (53.9%) | 52,592 (49.9%) | 2,216 (36.4%) |  |
| 40 to 59 | 15,147 (24.0%) | 9,519 (31.4%) | 6,682 (30.2%) | 35,093 (33.3%) | 1,779 (29.3%) |  |
| 60 and above | 7,738 (12.3%) | 8,078 (26.6%) | 3,522 (15.9%) | 17,731 (16.8%) | 2,087 (34.3%) |  |
| **Nationality** |  |  |  |  |  | <0.001 |
| Malaysian | 45,912 (72.8%) | 29,844 (98.3%) | 22,090 (99.8%) | 102,244 (97.0%) | 6,057 (99.6%) |  |
| Non-Malaysian | 17,186 (27.2%) | 506 (1.7%) | 35 (0.2%) | 3,172 (3.0%) | 25 (0.4%) |  |
| **Sex** |  |  |  |  |  | <0.001 |
| Male | 38,348 (60.8%) | 14,402 (47.5%) | 11,431 (51.7%) | 54,395 (51.6%) | 3,386 (55.7%) |  |
| Female | 24,750 (39.2%) | 15,948 (52.5%) | 10,694 (48.3%) | 51,021 (48.4%) | 2,696 (44.3%) |  |
| **Presence of Comorbidities** |  |  |  |  |  | <0.001 |
| Yes | 19,450 (30.8%) | 6,667 (22.0%) | 4,658 (21.1%) | 11,394 (10.8%) | 1,229 (20.2%) |  |
| No | 43,648 (69.2%) | 23,683 (78.0%) | 17,467 (78.9%) | 94,022 (89.2%) | 4,853 (79.8%) |  |

Abbreviations: COVID-19, coronavirus disease; ICU, intensive care unit; VE, vaccine effectiveness; BNT162b2, Pfizer-BioNTech; CoronaVac, Sinovac.
Breakdown by states of residence have been omitted for brevity.
Percentages in parentheses are by vaccination type and timing.
The ‘Late’ group are individuals vaccinated in July to August 2021 (1 to 2 months post-vaccination during the outcomes observation period in September 2021), ‘Early’ group are individuals vaccinated in April to June 2021 (3 to 5 months post-vaccination).

Table 4 shows the incidence rates for the outcomes of interest (ICU admission and deaths) and the corresponding vaccine effectiveness estimates by vaccination status, type, and timing of vaccination. The incidence rates and effectiveness estimates are further stratified by age groups (18 to 39 years, 40 to 59 years, and 60 years and above).

**Table 4.**
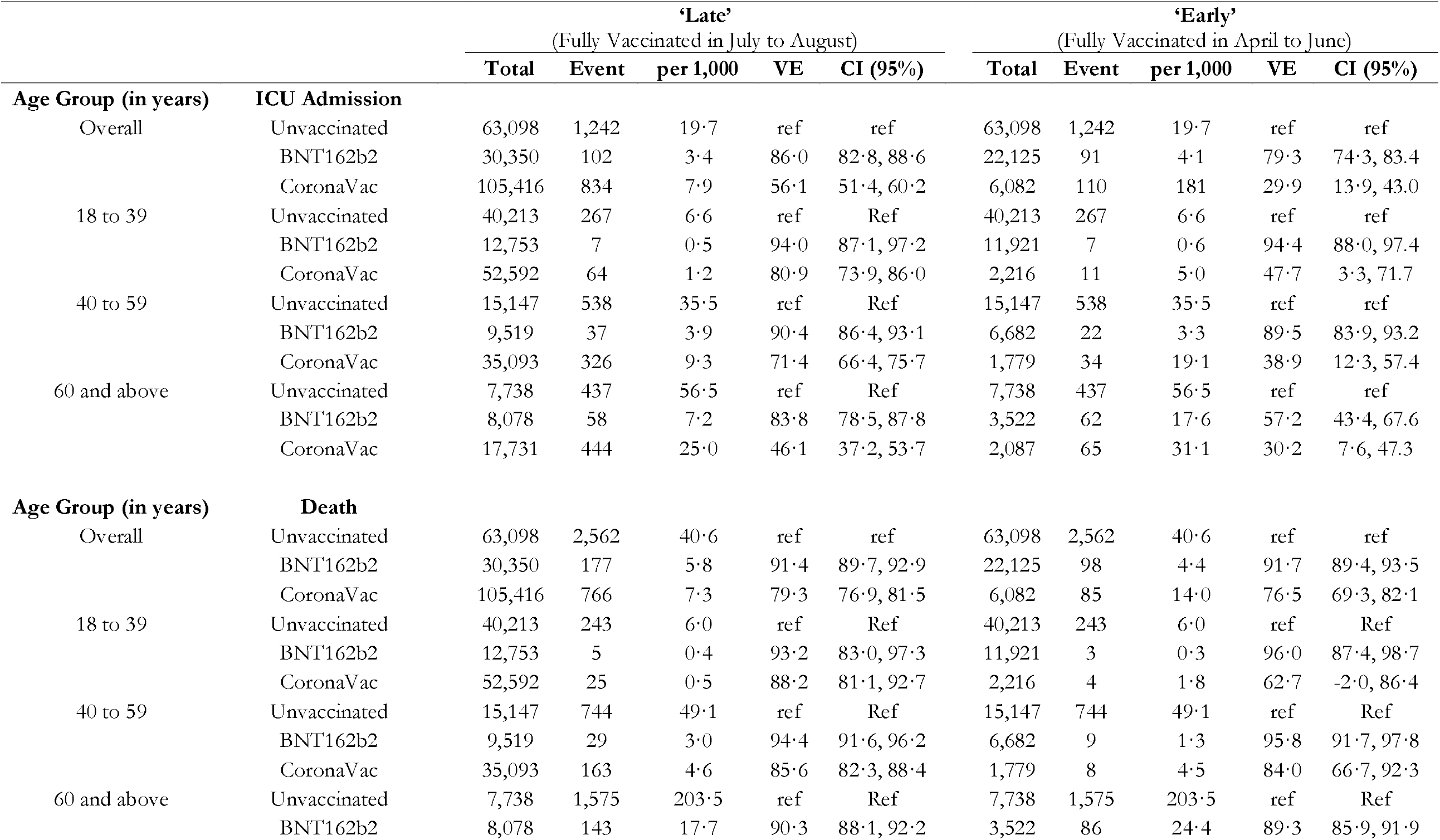
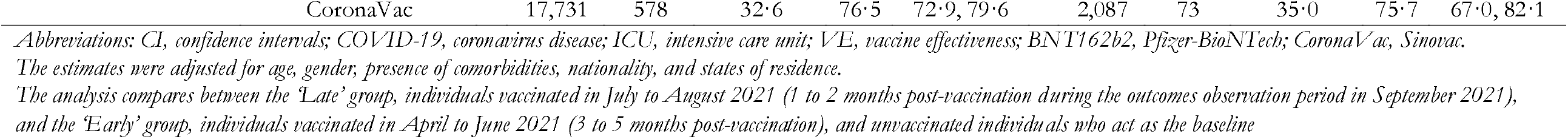
Vaccine effectiveness against COVID-19-related ICU admission and death by vaccine type and duration since full vaccination.

For BNT162b2, overall vaccine effectiveness against ICU admission and death were largely similar between the late and early groups. Overall vaccine effectiveness against ICU admissions dropped slightly from 86·0% (95% CI 82·8, 88·6) in the late group to 79·3% (95% CI 74·3, 83·4) in the early group. When stratified by age groups, the decline was stronger among cases aged 60 years and above, for which effectiveness fell to 57·2% (95% CI 43·4, 67·6). For deaths, vaccine effectiveness among both early and late groups remained at around 90%, suggesting no waning of effectiveness. When stratified by age, effectiveness against death among cases aged 60 years and above was lower than the other age groups, but similarly did not wane.

For CoronaVac, vaccine effectiveness waned for ICU admissions, but remained stable against deaths. Moreover, CoronaVac’s effectiveness estimates were generally lower compared to BNT162b2. For ICU admissions, vaccine effectiveness declined from 56·1% (95% CI 51·4, 60·2) in the late group to 29·9% (95% CI 13·9, 43·0) in the early group. The waning of effectiveness against ICU admissions was observed in all age groups. In contrast, vaccine effectiveness against death remained at above 75%. The age-stratified analysis shows that CoronaVac’s effectiveness against death remained stable for the older age groups (40 to 59 years old, 60 years and above), while for those aged 18 to 39 years old, the number of deaths were too small to estimate reliably.

Table S2 in the supplementary appendix shows that the findings are robust to redefining ICU admission and death as a single outcome. BNT162b2, consistent with the main analysis, showed no waning of effectiveness. CoronaVac showed waning effectiveness and lies between that of ICU admission and death. Figure S4 further shows the distribution of vaccine effectiveness estimated, by cycling through all combinations of covariates, against COVID-related ICU admission and death for by timing of vaccination for both vaccine types. While the unadjusted and partially adjusted effectiveness estimates vary in quantum from the fully adjusted estimates presented in Table 2, the broad findings are robust to the inclusion and exclusion of covariates.

## DISCUSSION

Overall, our study findings suggest significant waning of vaccine effectiveness against COVID-19 infections for both BNT162b2 and CoronaVac. Moreover, for CoronaVac, we found evidence of waning vaccine effectiveness against COVID-19-related ICU admissions but not for COVID-19 related deaths. The decline in vaccine effectiveness against COVID-19 infections was in accordance with trends observed in previous studies,^9–12^ but the decline in effectiveness among ICU admission warrants critical deliberation and, to the best of our knowledge, is among the first evidence of its kind for CoronaVac.

The magnitude of waning vaccine effectiveness against infection was observed to be greater among older recipients, particularly those aged 60 years old and above, in this retrospective population cohort. For BNT162b2, although effectiveness declined, it remained above WHO’s 50% primary efficacy endpoint.^7^ In contrast, our study observed significant declines in CoronaVac’s vaccine effectiveness against COVID-19 infections, particularly in the older age groups (40 to 59 years old, 60 years and above), among whom the COVID-19 infection incidence rates after three to five months of being fully vaccinated were similar to that of the unvaccinated.

We then proceeded to estimate the vaccine effectiveness for severe outcomes in a retrospective cohort of confirmed COVID-19 cases. Overall, we found that BNT162b2 vaccine remained highly protective against ICU admission and deaths over time. Studies elsewhere also demonstrated that BNT162b2 retained high effectiveness rates against severe outcomes for up to six months.^9,10^ There is, however, a small decline in effectiveness over time against ICU admission among individuals 60 years and above in our study. This is in line with findings in the US where effectiveness of BNT162b2 against hospitalisation declined slightly after 120 days for the elderly.^19^

For CoronaVac vaccine recipients, our study observed significant declines in vaccine effectiveness against ICU admission. Older age groups also registered lower effectiveness. This could be partly due to greater use of CoronaVac than BNT162b2 vaccines in the older and comorbid population who are at the greatest risk of developing severe illness, and hence prioritised in the early phases of PICK amid vaccine supply constraints,^20^ which our analysis attempted to control for. The lower effectiveness observed among older age groups is consistent with emerging evidence that older persons mount a lower immune response after a standard primary series of the CoronaVac vaccine compared to younger individuals and BNT162b2 recipients.^21,22^ This lower immune response was similarly observed among recipients who were immunocompromised when compared to healthy adult recipients.^23^ Findings from a study in Hong Kong SAR concluded that vaccination with BNT162b2 induced stronger humoral responses when compared to CoronaVac.^24^ CoronaVac’s neutralizing antibody levels have also been reported to fall by seven-fold within six months of vaccination.^24,25^ In addition, low effectiveness may also be attributed to the immunologic escape in the CoronaVac recipients against the Delta variant as the neutralisation property of this inactivated vaccine reduced by 31.6-fold compared to ancestral lineage, whilst the BNT162b2 was reduced by only three to five-fold.^26,27^

Although CoronaVac’s effectiveness against ICU admissions declined over time, our study showed that it retained substantial protection against death. Early reception of ICU care may have contributed towards better survival. In Malaysia, daily COVID-19-related ICU occupancy eased from an average of 1400 in July to August 2021 to an average of 1166 in September 2021.^16^ This allowed more COVID-19 cases to receive ICU care when necessary. Nevertheless, the waning of effectiveness against ICU admissions after three to five months of full vaccination with the CoronaVac vaccine warrants concern and motivates consideration for the timing and need for booster dose(s) among its recipients. The lower vaccine effectiveness and subsequent waning suggests that CoronaVac primary vaccination series may require three doses, as the protection from the current two-dose regime appears inadequate, as per WHO recommendation.^22^ In Singapore, recipients of the CoronaVac vaccine are now required to complete a three-dose primary vaccination series.^28^

Our study has important policy implications. In Malaysia, at the point of writing, boosters are offered to all CoronaVac vaccine recipients who had been fully vaccinated for at least three months prior, consistent with our findings that CoronaVac’s vaccine effectiveness significantly waned after at least three months. On the other hand, recipients of the BNT162b2 vaccine were initially offered boosters after six months of being fully vaccinated, before being reduced to three months beginning 28 December 2021. The rollout of booster doses prioritised the elderly (60 years and above) before younger age groups in age de-escalation manner. This follows our finding that waning of effectiveness was observed more strongly among older age groups. The narrative here highlights the rollout of booster doses in Malaysia as an example of evidence-based policymaking.

Our findings on waning effectiveness for CoronaVac may guide policies in LMICs that utilised CoronaVac in respective rollouts. The decline in effectiveness against ICU admissions, especially among individuals aged 60 years and above, signals caution towards its continued usage contingent on country-specific context. In Malaysia, due to vaccine inequity and supply challenges, CoronaVac was used in all individuals aged 18 years and above, including the elderly, in the early phases of PICK. When data emerged that the effectiveness of CoronaVac against ICU admissions waned, booster doses were offered promptly. Nevertheless, due to its continued protection against deaths, CoronaVac was retained in PICK. For policymakers in LMICs, future use of the CoronaVac vaccine should consider carefully its waning effectiveness over time, especially against ICU admission. Any targeted rollouts of CoronaVac should further consider its age-specific protection over time.

Our study has three strengths. First, we utilised a rich consolidated database from multiple official and granular data sources, which are nationally representative of Malaysia. Second, by restricting the outcome observation period to September 2021 when lockdown measures were harmonised nationwide and when testing remained high, we were able to estimate vaccine effectiveness and compare the different timing of vaccination with minimal potential unobserved confounders. Third, our findings on CoronaVac’s effectiveness over time, to our best knowledge, is among the first investigated and made available. This may have important implications on the future monitoring of this vaccine, as it is one of the most widely used vaccine worldwide.^14^

However, our study is not without limitations. First, in the absence of a national testing strategy during the study period (September 2021), the capturing of COVID-19 infections may not be standardised nationally, although testing remained high. Second, the lack of adequate genomic surveillance in Malaysia rendered impossible to ascertain variant-specific vaccine effectiveness, and their variation, if any, over time. Our study period also preceded the detection of the Omicron variant of concern.

Moving forward, we recommend that the monitoring of vaccine effectiveness over time remains prioritised to guide policy decisions on the rollout of booster dose(s). To build on our study findings, monitoring of effectiveness for other vaccine platforms used in Malaysia and among adolescents, which were first offered the COVID-19 vaccines in September 2021, should be initiated. Genomic surveillance efforts should also be strengthened to enable investigation into variant-specific vaccine effectiveness. Finally, as booster doses are rolled out, the performance and safety of the booster doses should be assessed to furnish data needs for policy calibration. In several studies conducted elsewhere, early findings on booster doses have been encouraging.^29,30^

## Data Availability

The data collected for the study, including de-identified participant data will be available upon request

## Authors Contribution

JLS, MH, PSKT, BHT, TT, EVL, KMP and SS designed the study. MRA, HY, SMZ, and FMZ collected the data. JLS, MH and TT performed the data curation, analysis and visualization. The data was interpreted by all authors. PSKT wrote the original draft of the manuscript. All authors reviewed and edited the manuscript. JLS, MH and TT accessed and verified the data. The study was supervised by BHT, KMP, and SS. All authors had full access to all data in the studies and had final responsibility for the decision to submit for publication.

## Conflict of interests

The authors declare that they have no conflict of interests.

## Acknowledgements

We would like to thank the Director-General of Health Malaysia for his permission to publish this article, as well as Dr Chee Peng Hor from the Ministry of Health Malaysia for his valuable feedback and comments on the article.

## Funding

No funding

## Notes

### Competing Interest Statement

The authors have declared no competing interest.

### Funding Statement

This study did not receive any funding

### Author Declarations

RECoVaM study and Ethical Consideration This study, commissioned by the Ministry of Health, Malaysia, is part of The Real-World Evaluation of COVID-19 Vaccines under the Malaysia National COVID-19 Immunisation Programme (RECoVaM) study registered in the National Medical Research Register (NMRR-21-1660-60697). This study was conducted according to guidelines in the Declaration of Helsinki and was granted ethnical approval by the Medical Research and Ethics Committee (MREC), Ministry of Health, Malaysia.

