## Supplemental File for "Waning COVID-19 Vaccine Effectiveness for BNT162b2 and CoronaVac in Malaysia: An Observational Study"

**Supplementary Appendix**

|  | |  | | **Table S1: Breakdown of overall utilisation by vaccine type in the *Program Imunisasi COVID-19 Kebangsaan* (PICK)** | | | | |
| --- | --- | --- | --- | --- | --- | --- | --- | --- |
|  | **Total (1 April 2021 to 31 August 2021)** | | | | **1 April 2021 to 30 June 2021** | | **1 July 2021 to 31 August 2021** | |
|  | **N** | | **%** | | **N** | **%** | **N** | **%** |
| **Total** | 15,083,858 | |  | | 2,121,393 |  | 12,962,465 |  |
| **BNT162b2** | 6,613,388 | | **43·8** | | 1,409,146 | **66·4** | 5,204,242 | **40·1** |
| **CoronaVac** | 7,636,740 | | **50·6** | | 711,190 | **33·5** | 6,925,550 | **53·4** |
| **AZD1222** | 803,578 | | **5·3** | | 65 | **<0·1** | 803,513 | **6·2** |
| **Others** | 30,152 | | **0·2** | | 992 | **<0·1** | 29,160 | **0·2** |

*Abbreviations: BNT162b2, Pfizer-BioNTech; CoronaVac, Sinovac; AZD1222, AstraZeneca
Vaccines in the ‘Others’ category include primarily Ad5-nCoV (CanSino), and BBIBP-CorV (Sinopharm)*

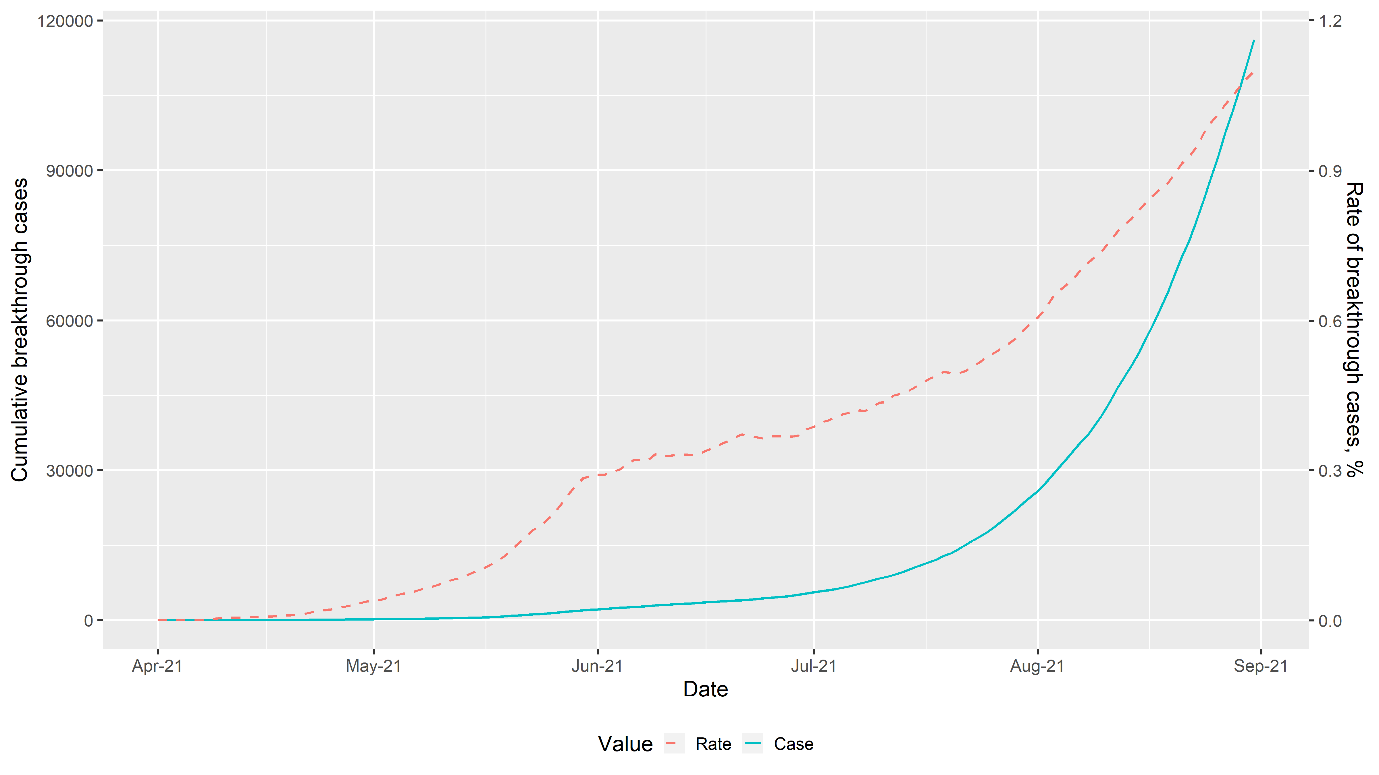

**Figure S1: Breakthrough level and rates for confirmed COVID-19 infection**

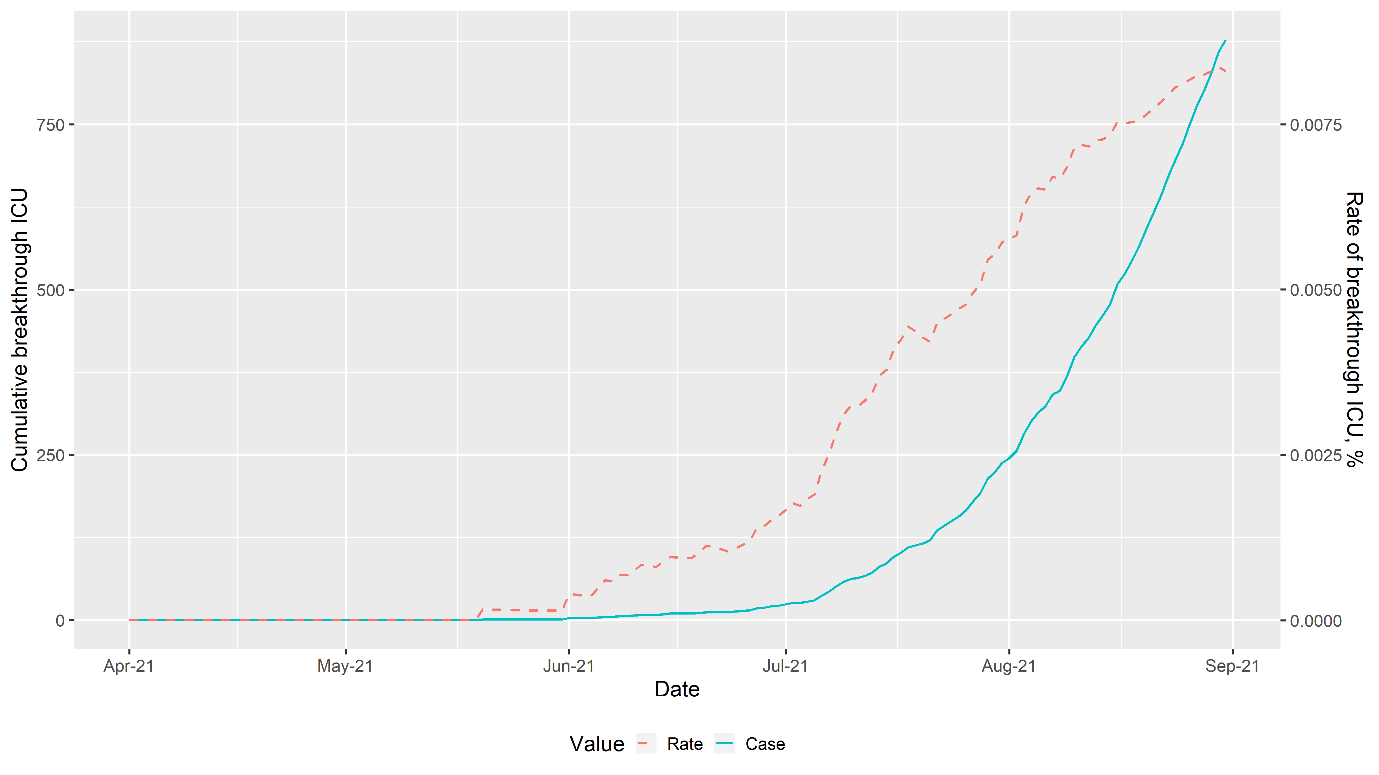

**Figure S2: Breakthrough level and rates for COVID-19-related ICU admissions**

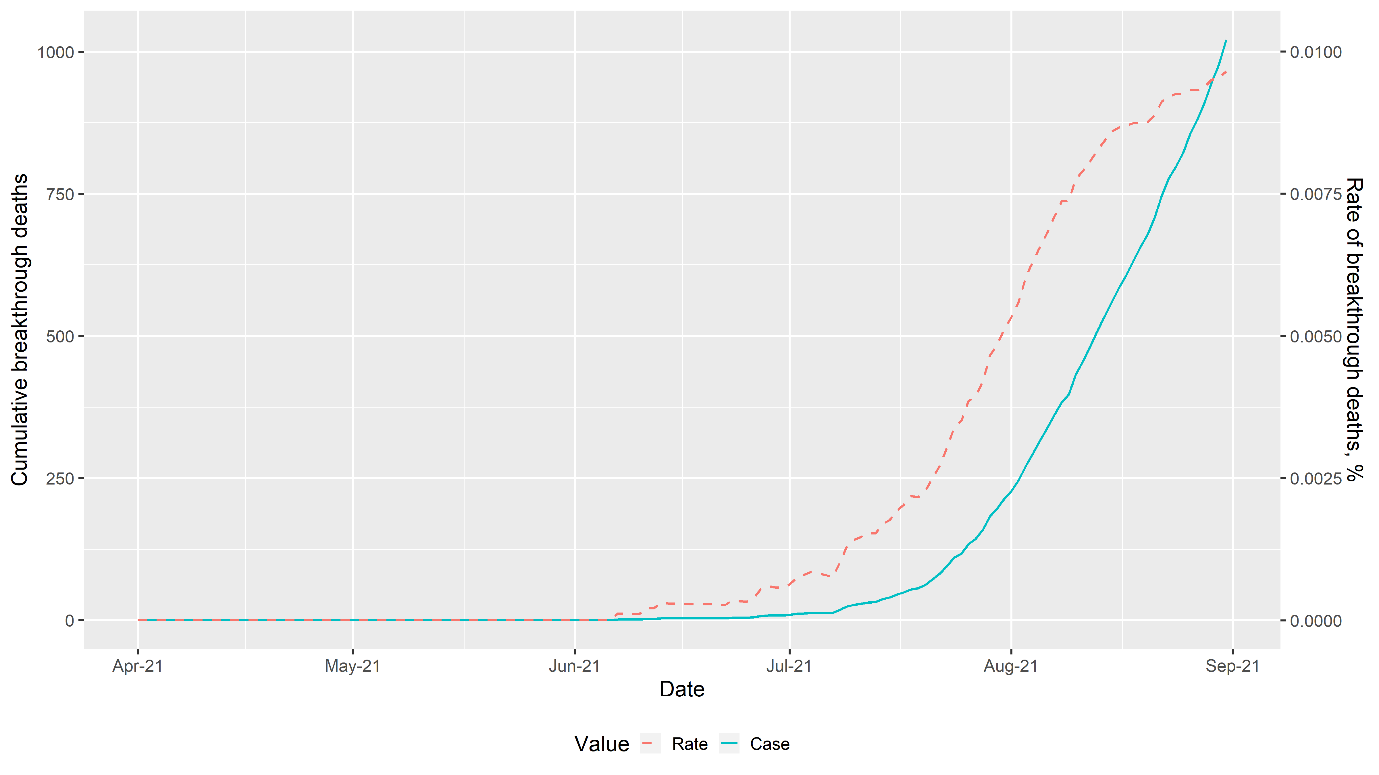

**Figure S3: Breakthrough level and rates for deaths due to COVID-19**

*Retrospective Population Cohort: Negative Binomial Regression*

Using aggregated count of outcomes (confirmed SARS-CoV-2 infection, COVID-19-related ICU admission, and death due to COVID-19), stratified by age group, sex, and state or region, this empirical strategy estimates the adjusted relative risk (RR) of outcomes for combinations of vaccination type and timing. The Klang Valley states (Kuala Lumpur, Putrajaya, and Selangor) are agglomerated into one single unit to reflect porous borders, especially for work purposes, throughout the COVID-19 epidemic. Moreover, the northern states (Perlis, Kedah, and Penang) are agglomerated into one unit due to cross-state vaccination, which may overstate vaccine coverage in select states. This is not an issue for the logistic regression approach described later, which uses individual-level variation in outcomes, rather than group-level variation. The age group variable is split into three levels – (i) aged between 15 to 39, (ii) aged between 40 to 59, and (iii) aged 60 and above. The lower bound, 15 years of age, reflects the age group intervals for which population data is available, with the offset being the person-days for the respective group.

$$\ln\left( Y \right)=\alpha+\left\{ \boldsymbol{Vax} \right\}\boldsymbol{\beta}+\boldsymbol{X\gamma}+\ln\left( Offset \right)+\epsilon$$

$$\left\{ {VE}_{Y} \right\}_{vt}=100*\left( 1-\exp\left( \beta_{vt} \right) \right)$$

We regress the count of the outcome of interest *Y* on a constant, and its stratification levels – (i) {*Vax*}, a vector containing the dummies indicating the vaccination type and timing, e.g., ‘early Sinovac’ group, (ii) ***X***, a vector containing the baseline characteristics, i.e., age groups, sex, and state, and (iii) *Offset*, the person-days, with its coefficient fixed to 1. Vaccine effectiveness is then calculated from the adjusted relative risk on the {*Vax*} vector. This produces a vector of vaccine effectiveness for each type and timing combination, from which the presence, and scale of waning vaccine effectiveness can be inferred.

*Retrospective Cohort of Confirmed COVID-19 Cases: Logistic Regression*

Amongst confirmed COVID-19 cases, this empirical strategy estimates the adjusted odds ratio using individual-level data on outcomes (COVID-19-related ICU admission, and death due to COVID-19), vaccination status and timing, as well as demographics for combinations of vaccination type and timing. Like the aggregate analysis described above, the Klang Valley states are agglomerated into one single unit, but the northern states are treated as separate units. This strategy adjusts for (i) age as a continuous variable, (ii) presence of comorbidities, (iii) sex, (iv) nationality, and (v) state fixed effects. As per an earlier study on Malaysia [9], the restriction of the cohort to only confirmed COVID-19 cases reflect the absence of individual-level data on demographics on individuals not infected by COVID-19. The vaccine effectiveness estimates, therefore, are interpreted as the risk reduction, according to vaccination type and timing, assuming infected in September 2021.

$$\Pr\left( Y_{i} \right)=\Lambda\left( \alpha+\left\{ \boldsymbol{Va}\boldsymbol{x}_{\boldsymbol{i}} \right\}\boldsymbol{\beta}+\boldsymbol{X}_{\boldsymbol{i}}\boldsymbol{\gamma}+\epsilon_{i} \right)$$

$$\left\{ {VE}_{Y} \right\}_{vt}=100*\left( 1-\exp\left( \beta_{vt} \right) \right)$$

We regress a dummy indicating if individual *i* experienced the outcome of interest *Y* on a (i) constant, (ii) **{*Vax*}**, a vector indicating the individual’s vaccination type and timing, and (iii) ***X***, a vector containing the adjustment factors. From the adjusted odds ratio on **{*Vax*}**, we calculate the effectiveness for each vaccine type and timing to evaluate the presence, and scale of waning effectiveness.

| **Table S2. Vaccine effectiveness against COVID-19-related severe outcomes (ICU admission or death) by vaccine type and duration since full vaccination** | | | | | | | | | | | | | |
| --- | --- | --- | --- | --- | --- | --- | --- | --- | --- | --- | --- | --- | --- |
|  |  |  | **‘Late’** (Fully Vaccinated in July to August) | | | | |  | **‘Early’** (Fully Vaccinated in April to June) | | | | |
|  |  |  | **Total** | **Event** | **per 1,000** | **VE** | **CI (95%)** |  | **Total** | **Event** | **per 1,000** | **VE** | **CI (95%)** |
| **Age Groups (in years)** | **Severe outcomes**  **(ICU Admission or Death)** |  |  |  |  |  |  |  |  |  |  |  |  |
| Overall | Unvaccinated |  | 63,098 | 3,120 | 49.4 | ref | ref |  | 63,098 | 3,120 | 49.4 | ref | ref |
|  | BNT162b2 |  | 30,350 | 224 | 7.4 | 90.7 | 89.2, 92 |  | 22,125 | 155 | 7.0 | 88.2 | 85.9, 90.2 |
|  | CoronaVac |  | 105,416 | 1,313 | 12.5 | 72.4 | 70, 74.5 |  | 6,082 | 159 | 26.1 | 63.4 | 56, 69.6 |
| 18 to 39 | Unvaccinated |  | 40,213 | 404 | 10.0 | ref | ref |  | 40,213 | 404 | 10.0 | ref | ref |
|  | BNT162b2 |  | 12,753 | 9 | 0.7 | 93.9 | 88.1, 96.9 |  | 11,921 | 7 | 0.6 | 95.6 | 90.7, 97.9 |
|  | CoronaVac |  | 52,592 | 81 | 1.5 | 81.2 | 75.3, 85.7 |  | 2,216 | 13 | 5.9 | 50.1 | 12.1, 71.6 |
| 40 to 59 | Unvaccinated |  | 15,147 | 1,000 | 66.0 | ref | ref |  | 15,147 | 1,000 | 66.0 | ref | ref |
|  | BNT162b2 |  | 9,519 | 48 | 5.0 | 93.4 | 91.1, 95.1 |  | 6,682 | 30 | 4.5 | 91.6 | 87.7, 94.2 |
|  | CoronaVac |  | 35,093 | 397 | 11.3 | 78.9 | 75.7, 81.6 |  | 1,779 | 37 | 20.8 | 58.2 | 40.9, 70.4 |
| 60 and above | Unvaccinated |  | 7,738 | 1,716 | 221.8 | ref | ref |  | 7,738 | 1,716 | 221.8 | ref | ref |
|  | BNT162b2 |  | 8,078 | 167 | 20.7 | 89.7 | 87.6, 91.5 |  | 3,522 | 118 | 33.5 | 84.9 | 81.1, 87.9 |
|  | CoronaVac |  | 17,731 | 835 | 47.1 | 69.9 | 66.3, 73.2 |  | 2,087 | 109 | 52.2 | 68.0 | 59.5, 74.8 |

*Abbreviations: CI, confidence intervals; COVID-19, coronavirus disease; ICU, intensive care unit; VE, vaccine effectiveness; BNT162b2, Pfizer-BioNTech; CoronaVac, Sinovac.*

*The estimates were adjusted for age, gender, presence of comorbidities, nationality, and states.*

*The analysis compares between individuals vaccinated in July to August 2021 (1 to 2 months post-vaccination during the outcomes observation period in September 2021), individuals vaccinated in April to June 2021 (3 to 5 months post-vaccination), and unvaccinated individuals who act as the baseline.*

| 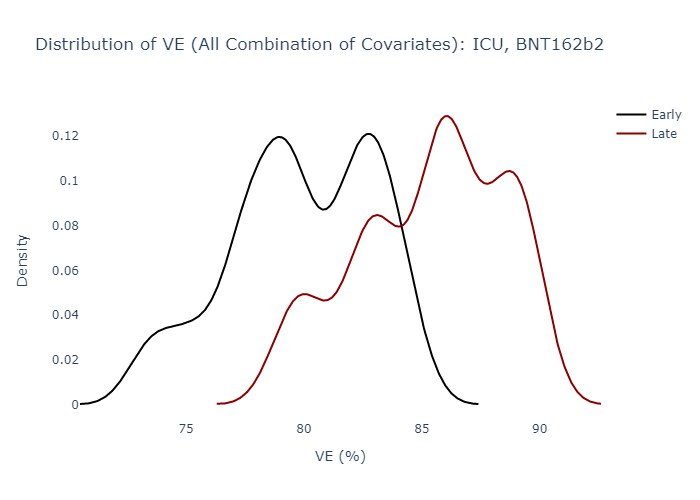  **(a)** | 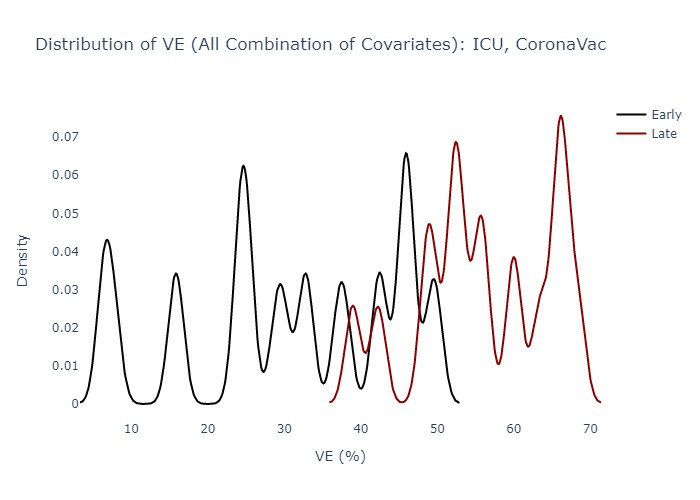  **(b)** |
| --- | --- |
| 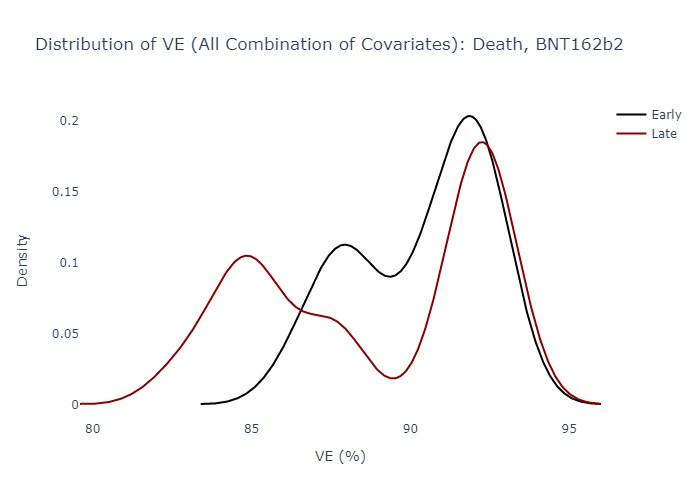  **(c)** | 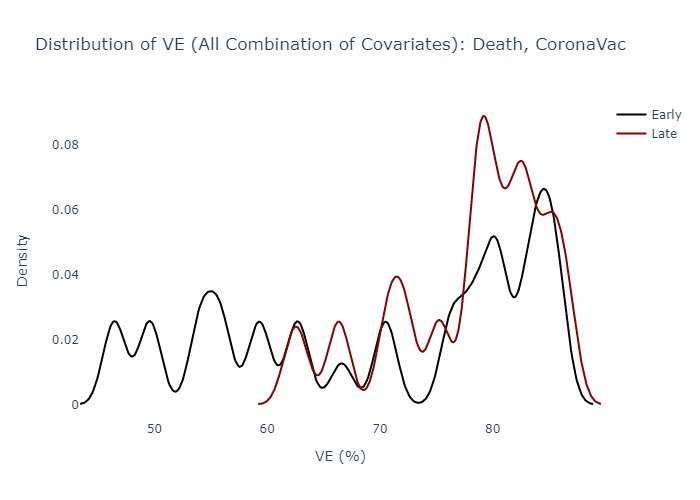  **(d)** |

**Figure S4: Distribution of vaccine effectiveness estimated with all permutations of covariates in the retrospective cohort of confirmed COVID-19 cases for BNT162b2 and CoronaVac by period of vaccination, and for COVID-19-related ICU admission and death**

*The analysis compares between individuals vaccinated in July to August 2021 (1 to 2 months post-vaccination during the outcomes observation period in September 2021), individuals vaccinated in April to June 2021 (3 to 5 months post-vaccination), and unvaccinated individuals who act as the baseline.*
